# Exploration of interethnic variation in the ibuprofen metabolizing enzyme CYP2C9: a genetic-based cautionary guide for treatment of COVID-19 symptoms

**DOI:** 10.1101/2021.01.09.21249508

**Authors:** Ammar Ali Almarzooq

## Abstract

Coronavirus disease 2019 (COVID-19), is a rapidly spreading infectious illness that causes a debilitating respiratory syndrome. While non-steroidal anti-inflammatory drugs (NSAIDs), may be prescribed for the management of pain and fever, there was early controversy on the use of ibuprofen for symptomatic treatment of COVID-19. P450 enzyme CYP2C9 are known to be involved in the metabolism of NSAIDs. Although no study has been conducted in the setting of population genetics in patients with COVID-19 yet, there are plausible mechanisms by which genetic determinants may play a role in adverse drug reactions (ADRs). In this work, we adjusted expected phenotype frequencies based on racial demographic models dependent on population ancestry in drug responses and toxicity events associated with ibuprofen treatment. A cohort of 101 Jordanian Arab samples retrospectively were selected and genotyped using Affymetrix DMET Plus Premier Package, within the context of over 100,000 global subjects in 417 published reports. European populations are 7.2x more likely to show impaired ibuprofen metabolism than Sub-Saharan populations, and 4.5x more likely than East Asian ancestry populations. Hence, a proactive assessment of the most likely gene-drug candidates will lead to more effective treatments and a better understanding of the role of pharmacogenetics for COVID-19.

## Introduction

The global pandemic of the novel coronavirus disease 2019 (COVID-19) has caused a global healthcare crisis resulting from high infection and mortality rates^1^. The National Health Commission of China released key guidelines for the diagnosis and treatment of pneumonitis, and related diseases, caused by COVID-19^2^. Such guidelines included oxygen therapy, mechanical ventilation and drug therapy, as well as provided recommendations for simultaneous drug treatments in extreme cases. For suspected or confirmed cases of COVID-19, requiring urgent care for conditions such as fever and/or sore throat, pharmacological management may require antibiotics and/or analgesics, an alternative^2^. Specifically, non-steroidal anti-inflammatory drugs (NSAIDs) e.g. ibuprofen, may be prescribed for the management of pain and fever. However, uncertainty related to infection etiology and efficacy, and emerging concerns related to the use of common NSAIDs,have presented additional challenges in the treatment of COVID-19. Fang and colleagues at the beginning of the pandemic hypothesized a possible deleterious role of ACE2-stimulating drugs and ibuprofen on the course of SARS-CoV-2 infected patients^3^. The increase of ACE2 expression may lead to a potential rise of SARS-CoV-2 viral load and consequently to a more severe disease course. Therefore, the authors concluded their commentary discouraging the use of these drugs in the setting of COVID-19. This plausible mechanism together with the above-reported evidences of ibuprofen, led the National Agency for the Safety of Medicines and Health Products (ANSM) of France to release a warning, asking whether patients showing symptoms of COVID-19 should use paracetamol rather than ibuprofen^4^. This warning was echoed by the British Medical Journal^5-7^, causing an 80% drop in ibuprofen prescriptions in France^8^. The UK Medicines and Healthcare products Regulatory Agency (MHRA) reported that, in the absence of clear evidence, patients should be advised to take paracetamol to treat the symptoms of COVID-19, unless paracetamol is not suitable for them^9^. However, the World Health Organization (WHO), after initially advising against the use of ibuprofen for COVID-19, quickly retracted the public^10^. Indeed, the scientific debate on ibuprofen use for COVID-19 continues^10-13^. Abu Esba and colleagues prospectively recruited 503 adults in Saudi Arabia with confirmed SARS-CoV2 infection of whom 40 (8%) used ibuprofen during the infection, 17 (3.4%) were assumed to have used other NSAIDs, and 96 (19%) were chronically treated with NSAIDs before and during the infection. Neither the acute nor the chronic use of NSAIDs resulted in increased mortality or severe COVID-19^14^. Also, Kragholm and colleagues reported the data of a retrospective, Danish-based cohort study, including 4002 adults with COVID-19 of whom 264 (6.6%) were treated with ibuprofen^15^. Again, no significant association between ibuprofen and severe COVID-19 was found. Finally, Rinott and colleagues retrospectively evaluated the use of ibuprofen versus paracetamol during the course of SARS-CoV2 infection in Israel across 403 adult patients, confirming that ibuprofen was not associated with severe COVID-19^16^. However, currently no study has been conducted in the setting of population genetics.

Genetic factors are one of the major contributors to individual or ethnic differences in drug therapeutic efficacy and toxicity^17, 18^. Consequently, host genetics and demography associated with COVID-19 are crucial aspects of infection and prognosis. Thus, integral medication dosing might need to be altered based on a patient’s genetic information^19, 20^. There are several gene variants that alter how an individual metabolizes and processes ibuprofen, potentially increasing the risk of undesirable ADRs. The cytochrome P450 enzyme CYP2C9, facilitates metabolism of several NSAIDs, including ibuprofen, and *CYP2C9* allele frequencies have been shown to vary substantially across diverse ethnic groups^21, 22^.

In March 2020, the Clinical Pharmacogenetics Implementation Consortium (CPIC) published a pharmacogenetic guideline on NSAIDs, with specific therapeutic recommendations based on *CYP2C9* phenotype^23^. The phenotype was derived from an activity score, obtained by the sum of two individual allele scores.Recent meta-analysis of *CYP2C9* alleles and ibuprofen concentrations using the Pharmacogenomics Knowledge Base (PharmGKB), showed strong correlations between the CYP2C9*2 allele (p.R144C; rs1799853) and *CYP2C9*3* allele (*p*.*I359L; rs1057910*) with plasma levels of ibuprofen^24-28^. Based on these results, PharmGKB assigned the highest level of evidence (level 1A) to these associations, indicating these biomarkers were extensively studied and validated in clinical practice.

However, the goal of this study was to demonstrate that current pharmacogenomics databases can be leveraged to enhance the identification of a gene alleles, and to determine population differences in associated drug response/toxicity events. Results from this work have wide ranging impacts on the targeted treatment of COVID-19 patients across broad geographic ranges and ethnic backgrounds, thereby facilitating the drug development processes and help with prescribing therapeutics more safely.

## RESULTS

### Selection and analysis of CYP2C9 Variations

Eighteen *CYP2C9* variants across 101 Jordanian individuals of Arab descent associated with reduced enzyme function were selected (Table 1 & Table S1). The defective allele *\*2 (rs1799853)* was the most abundant variant (0.094), followed by allele *\*3* (rs1057910) (0.084). In addition, two rare variants, *c*.*1425A>T (rs1057911)* and *50196C>T (rs2017319)*, were also detected. These two SNPs had frequencies of less than 0.005. *CYP2C9 *2* and *\*3* together accounted for 17.8% of the allele frequency and about 32.7% of the reduced or non-functional genotype/phenotype associations. The four genotype frequencies *CYP2C9 *1/*1, *1/*2, *1/*3* and *\*2/*3* were 0.673, 0.158, 0.139 and 0.03, respectively. Moreover, the genotype frequencies showed no deviation from HWE (p□>□0.05; Table 2).

**Table 1.** Details of the 18 allele frequencies of *CYP2C9* gene (*NG_008385*.*1*; *NP_000762*.*2*)

| SNP | *CYP2C9* | Marker | Minor Allele |
| --- | --- | --- | --- |
|  | Allele | Call Rate | Frequency |
| *g.8633C>T; p.(Arg144Cys)* | *2 | 100 | 0.094 |
| *g.47639A>C; p.(Ile359Leu)* | *3 | 99 | 0.084 |
| *g.47640T>C; p.(Ile359Thr)* | *4 | 99 | 0 |
| *g.47644C>G; p.(Asp360Glu)* | *5 | 100 | 0 |
| *g.15626delA; p.(Lys273fs)* | *6 | 100 | 0 |
| *g.5080C>A; p.(Leu19Ile)* | *7 | 99 | 0.198 |
| *g.15560A>G; p.(His251Arg)* | *9 | 100 | 0 |
| *g.15623A>G; p.(Glu272Gly)* | *10 | 100 | 0 |
| *g.47567C>T; p.(Arg335Trp)* | *11 | 100 | 0 |
| *g.55363C>T; p.(Pro489Ser)* | *12 | 100 | 0 |
| *g.8301T>C; p.(Leu90Pro)* | *13 | 100 | 0 |
| *g.8577G>A; p.(Arg125His)* | *14 | 100 | 0 |
| *g.14125C>A; p.(Ser162Ter)* | *15 | 100 | 0 |
| *g.38522A>G; p.(Thr299Ala)* | *16 | 100 | 0 |
| *g.8556_8565delAGAAATGGAA;*<br>*p.(Lys118fs)* | *25 | 100 | 0 |
| *g.55221C>T; p.(Ala441=)* | NA | 99 | 0.045 |
| *g.47637A>G; p.(Tyr358Cys)* | NA | 100 | 0 |
| *g.55323A>T; p.(Gly475=)* | NA | 100 | 0.045 |

**Table 2.** Distribution of CYP2C9 alleles and genotypes in Jordanian Arabs (NG_008385.1; NP_000762.2).

| Allele | Variant | Substitution | Allele Count | Allelic Frequency | Genotype | Genotypic Frequency | Observed Genotypes | Expected Genotypes | $\chi^2$ | *p*-value |
| --- | --- | --- | --- | --- | --- | --- | --- | --- | --- | --- |
| *2 | *g.8633C>T; p.(Arg144Cys)* | C | 183 | 0.91 | CC | 0.81 | 82 | 82.89 | 1.1 | 0.58 |
|  |  | T | 19 | 0.09 | CT | 0.19 | 19 | 17.21 |  |  |
|  |  |  |  |  | TT | 0.00 | 0 | 0.89 |  |  |
| *3 | *g.47639A>C; p.(Ile359Leu)* | A | 185 | 0.92 | AA | 0.83 | 84 | 84.72 | 0.9 | 0.65 |
|  |  | C | 17 | 0.08 | AC | 0.17 | 17 | 15.57 |  |  |
|  |  |  |  |  | CC | 0.00 | 0 | 0.00 |  |  |

Significant D’ values were observed spanning the entire genomic region, following LD measurements for pairs of SNPs distributed across the 52-kb region. Most allele pairs of significant D’ values were observed spanning the entire genomic region, following LD measurements for pairs of SNPs distributed across the 52-kb region. Most allele pairs of *CYP2C9* had a D′ value equal to 1.0 (indicating complete LD), whereas, r2 values across the same region, show a LD block between the *\*7* allele (*rs67807361*) at exon 1 and the *\*14* allele (*rs72558189*) at exon 3. A clear LD block was also observed between *CYP2C9*3* (*rs1057910*) at exon 3 and between c.1425A>T (*rs1057911*) at exon 9, crossing an approximately 8-kb region (Figure 1A and Table S2). The experimental group *rs67807361* was significantly different from the other sampled populations (p=4.9×10-22; Table S3). However, the nucleotide BLAT search showed that the DNA sequence obtained from the flanking region of this SNP (124bp) had 100% sequence identity with the *CYP2C19* gene at the region of 10:94762716-94762839 (Figure 1B). Therefore, this variant was excluded from the analyses since the individual probes in the MIP assay only bind to a genomic footprint of ∼40bp. Thus, the homologous sequences would likely result in false-positive or false-negative variant calls^29^.

**Figure 1:**
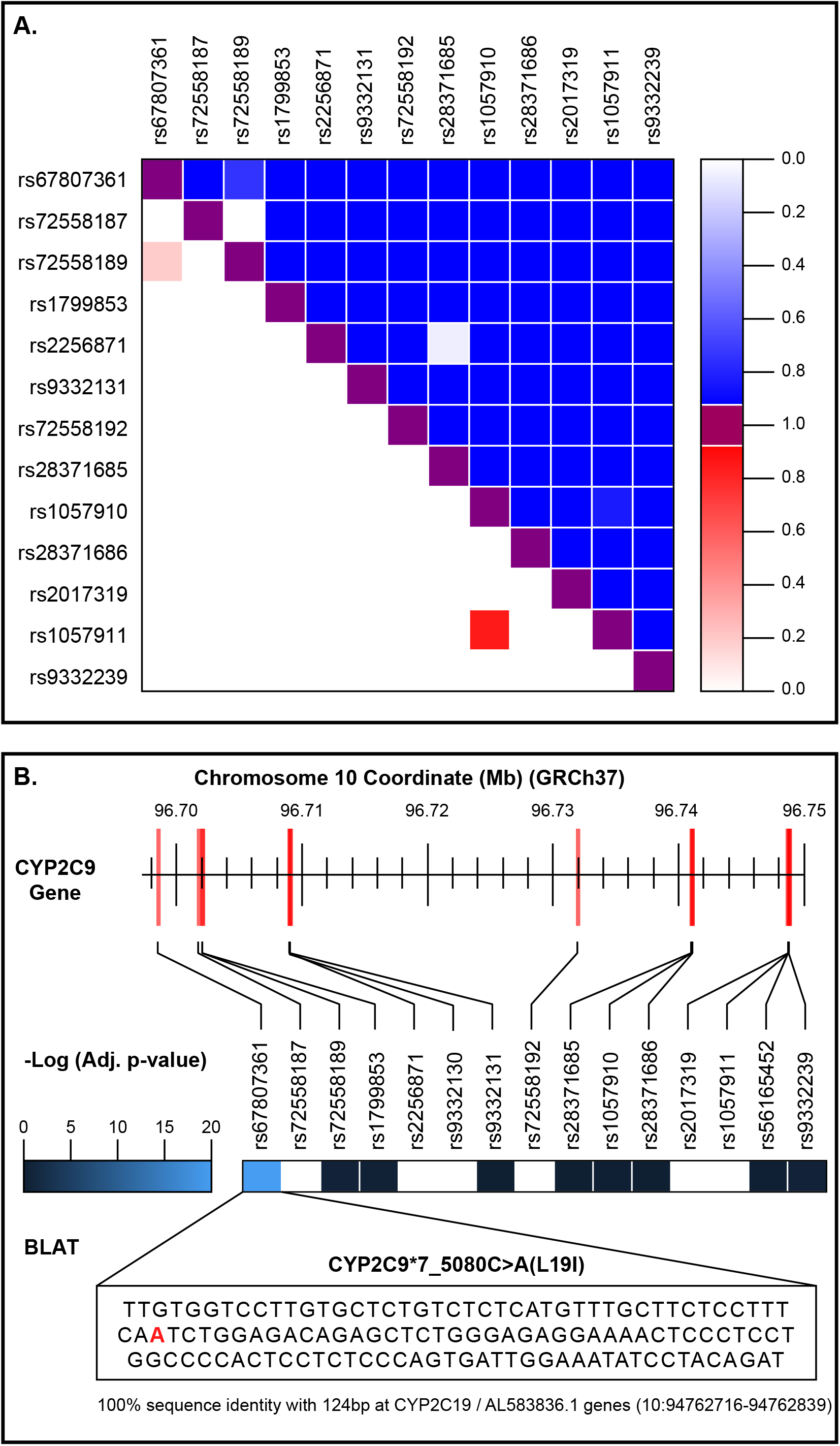
*CYP2C9* genetic interpretation in the context of Linkage Disequilibrium (LD) and high homology sequence. **a**. LD (D’ and r^2^) between 13 single nucleotide polymorphisms (SNPs) in the *CYP2C9* gene. **b**. Statistical comparison of allele frequencies on experimental data and reference populations. Color intensity indicates a more significant difference between the given experimental data and the population.

### Genetic structure of CYP2C9 across populations

The two leading principal components from the 13 variants shared between the Jordanian Arab population and the 22 global populations from the 1000 Genomes Project Phase III (1kG-p3) dataset (Figure 2A), captured 60.63% and 21.38% of the variance respectively, showing a well-defined separation between Jordanian Arabs and AFR, EAS, and SAS super populations. Jordanian Arabs had a close affinity with EUR (comprised of GBR, FIN, TSI, CEPH, and IBS), and validated by pairwise Fst analyses (Table S4). The lowest level of differentiation was observed between the Jordanian Arab population and GBR (Fst=5.97 × 10-3), followed by IBS (Fst=6.39 × 10-3), and FIN (Fst=6.69 × 10-3), whereas the greatest divergence was observed with GWD (Fst=8.54 × 10-2).

**Figure 2:**
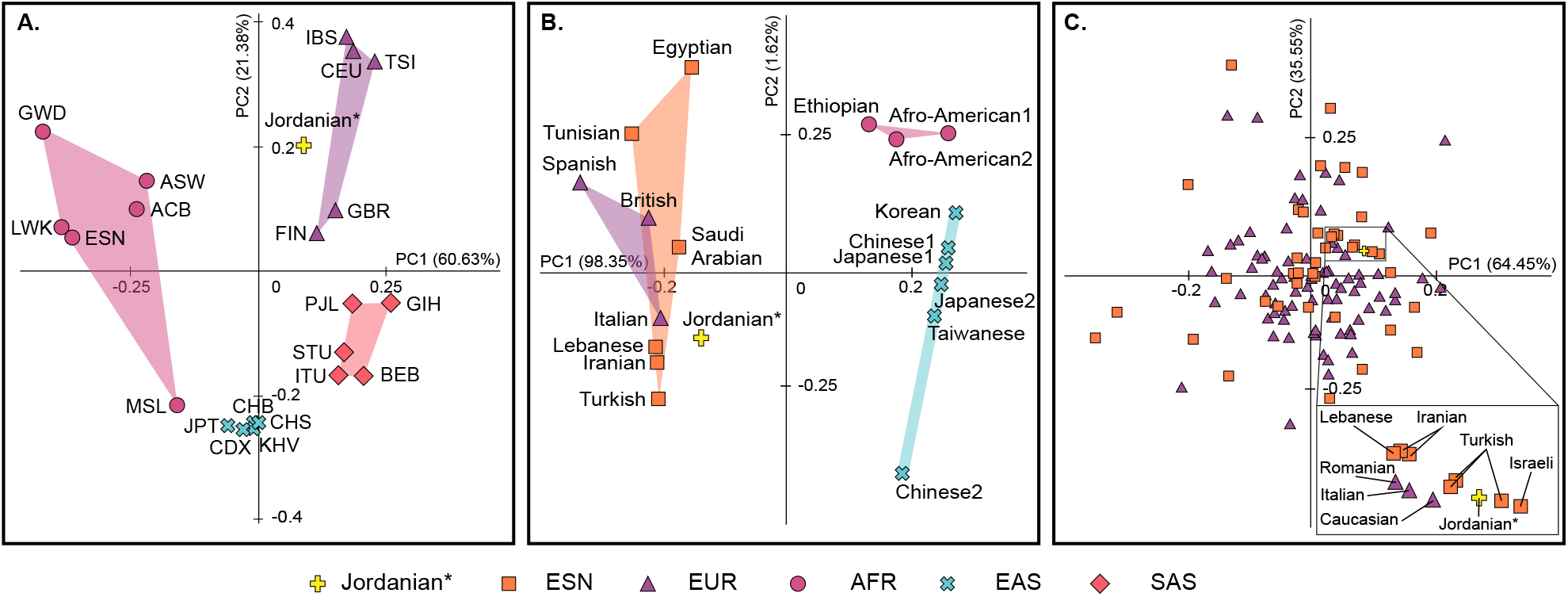
Population structure analysis of the *CYP2C9 *1, *2* and *\*3* alleles. **a**. MDS plot of the 14 variants shared between the Jordanian Arab population and 22 global populations from the 1000 Genomes Project Phase III (1kG-p3) dataset. **b**. MDS plot of the Jordanian Arab population and 18 public worldwide populations. **c**. MDS plot of the Jordanian Arab population and 118 populations from the Near Eastern (ESN) and European (EUR) groups.

Lack of genomic data for additional ethnic groups in the 1000 Genomes Project such as Middle Eastern populations (ESN), can reduce robustness and potentially result in biased geographic-based genomic analysis. Therefore, a secondary analysis was performed to include under-represented Arab populations. The two leading principal components shared between the Jordanian Arab population, and the 18 global reports including ESN for *\*1, *2* (*rs1799853*) and *\*3* (*rs1057910*); (Table S5) captured 98.35% and 1.62% of the variance, respectively, suggesting a well-defined genetic separation between Jordanian Arabs and AFR and EAS populations (Figure 2B). In addition, defined clusters of EUR and ESN populations were found, which were further validated using pairwise Fst analyses (Table S6). The lowest level of differentiation was observed between the Jordanian Arab population and Saudi Arabian population (Fst=7.4 x10-4), followed by Italian (Fst=1.62 × 10-3) and Turkish populations (Fst=1.8 × 10-3), whereas the greatest divergence was observed with the Korean population (Fst=1.13 × 10-1).

### Pharmacogenetic analyses by biogeographic grouping system

Across the nine biogeographical groups, 27% of subjects were of East Asian origin, followed by Europeans (26%), South Central Asians (13%), Near Easterns (12%), Americans (7%), Latinos (6%), African Americans/Afro-Caribbeans (4%), Sub-Saharan Africans (4%), and Oceanians (1%) (Table 3).Distinct differences were found among these populations, with direct impacts on ibuprofen clinical outcomes (Figure 3B). *The CYP2C9 *2* (*rs1799853*) and *\*3* (*rs1057910*) allele frequencies were significantly higher in the Central/South Asian origin (0.224), followed by Near Easterns (0.212), Europeans (0.203), Jordanian Arabs (0.178), and Latinos (0.116), indicating a decreased metabolism and clearance of ibuprofen as compared to Americans (0.064), Oceanians (0.045), East Asians (0.04), African Americans/Afro-Caribbeans (0.036) and Sub-Saharan Africans (0.024, Table S7). These significant variant alleles and genotypes were classified as PharmGKB Level 1A evidence with reduced enzyme function, and therefore are associated with recommended changes to ibuprofen dosing^22^. Interpretation of the translation into specific dosing guidelines for individual ibuprofen-diplotype pairs^23^ showed that Central/South Asian, Near Eastern, and European populations are 7.9x to 5.9x more likely to show impaired CYP2C9 metabolism than African populations (Sub-Saharan and African American/Afro-Caribbean populations, respectively), and 4.9x more likely than East Asian populations (Table S7).

**Table 3.**
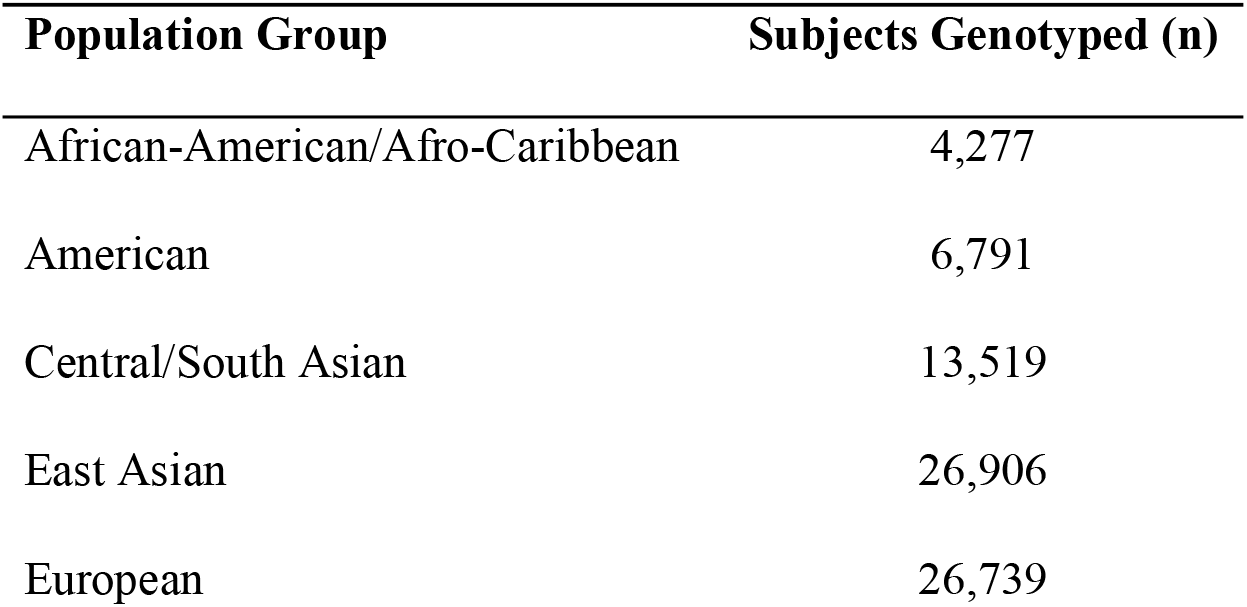
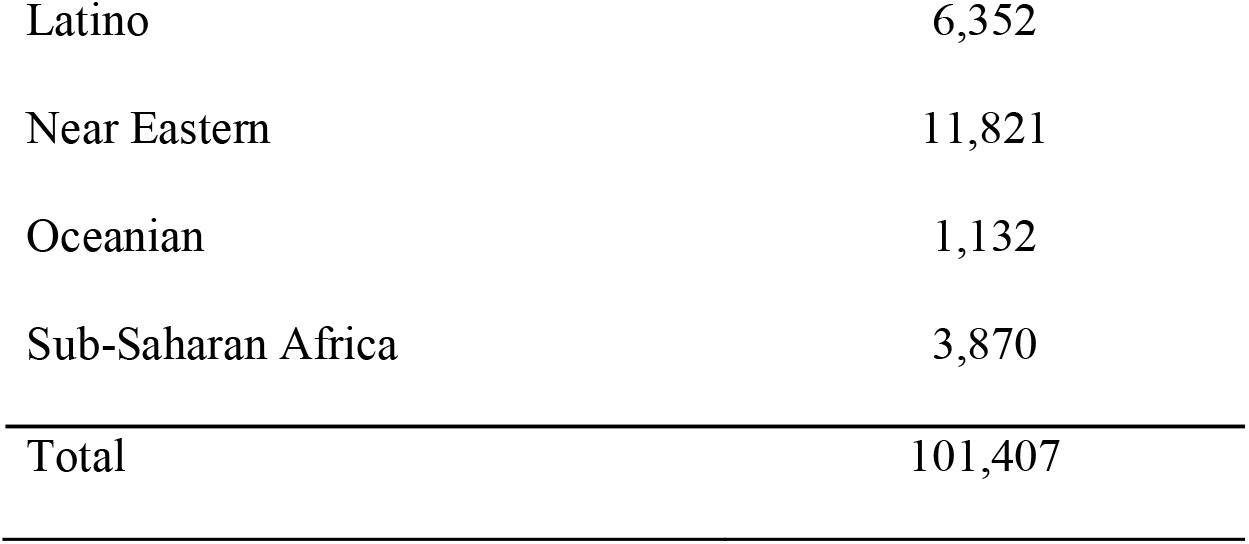
Sample size accumulated per biogeographical group

**Figure 3:**
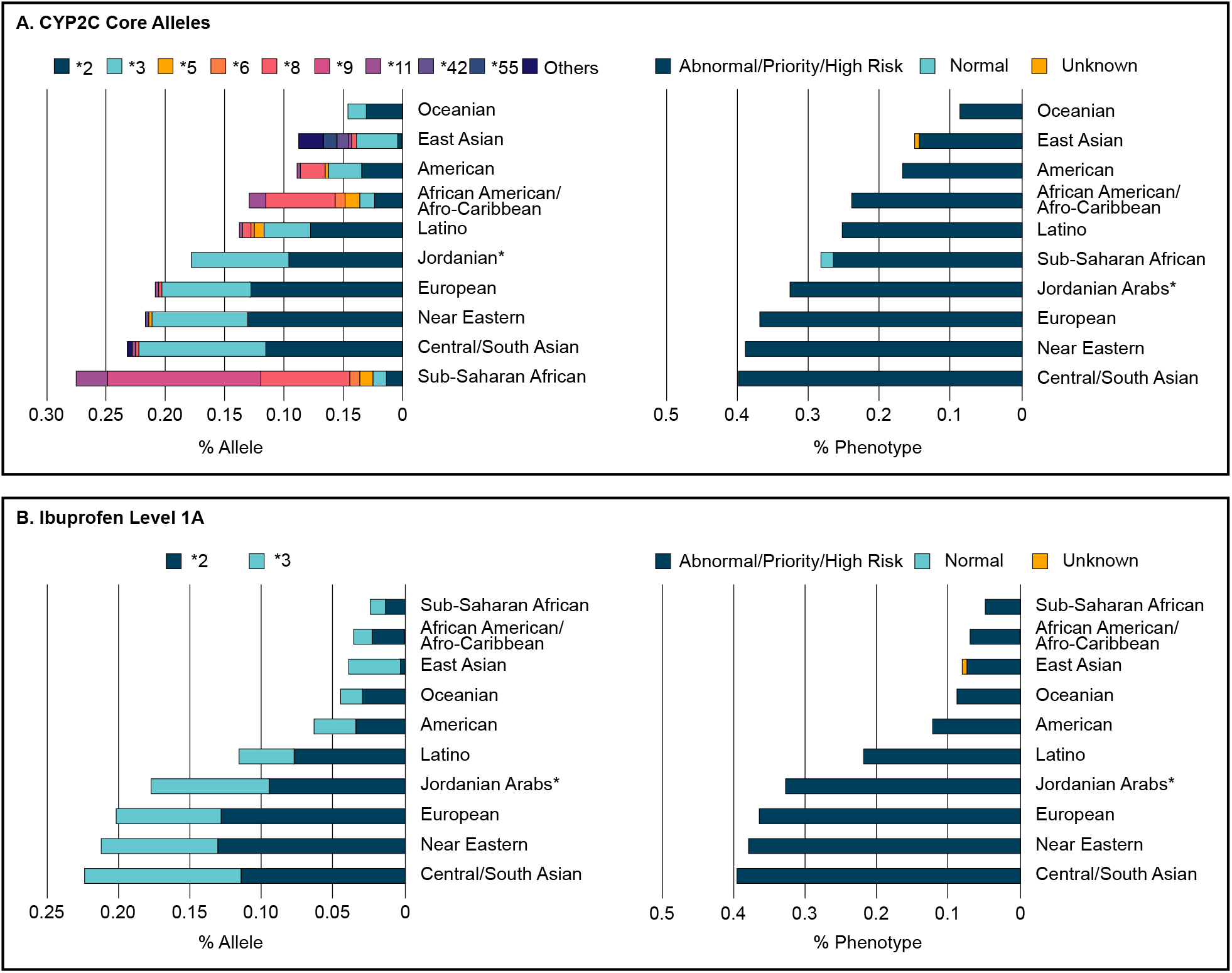
Biogeographical groups and pharmacogenetic analyses for CYP2C9 and ibuprofen associations. **a**. Frequencies of 13 actionable pharmacogenomics (PGx) alleles and phenotypes assessed cumulatively for the Jordanian Arab population against the nine biogeographical groups. **b**. Mapping of the total allele and phenotype frequency of the CYP2C9 alleles *\*2* and *\*3*.

Interestingly, a large number of generally less common alleles were also identified based on this approach (Table S7). Allele *\*9* (*rs2256871*) was significantly over-represented in Sub-Saharan Africans (0.13), but was not detected in other global populations (Figure 3A). Alleles *\*5* (*rs28371686*), *\*6* (*rs9332131*), *\*8* (*rs7900194*) and *\*11* (*rs28371685*) were significantly over-represented in African populations (Sub-Saharan African and African Americans/Afro-Caribbean) and under-represented in other populations. East Asian populations over-represented alleles *\*42* (*rs12414460*) and *\*55* (*42620C>A*).

## DISCUSSION

Although no direct evidence of pharmacogenomics data in patients with COVID-19 was available at the time of writing this manuscript, there are plausible mechanisms by which genetic determinants may play a role in adverse drug responses. While NSAIDs, including ibuprofen, may be prescribed for the management of pain and fever^2^, controversy arose on the use of ibuprofen due to the possibility of a worse COVID-19 prognosis^3-9^. Ibuprofen may offer symptomatic relief, and could provide healthcare professionals additional time to deliver customized care. Although this strategy is not always reliable since individuals may respond differently to similar treatments^22, 23^. In this work, several genetic markers were analyzed across diverse ethnic backgrounds to identify population differences in drug responses and toxicity events associated with ibuprofen treatment. Results from this study showed that pharmacogenomics studies can be leveraged to enhance the understanding of adverse reactions to the treatment of COVID-19 symptoms and support advancement of drug development pipelines.

MDS and Fst analyses showed that the Jordanian Arab population clustered with multiple regions within European and Near Eastern. These results validated that the current Jordanian population today falls into two main groups: one sharing more genetic characteristics with modern-day Europeans and Central Asians, and the other with closer genetic affinities to Arabia^30^. In addition, our analyses are in agreement with recent studies using large-scale genomics that indicated three major genetic events related to Levant populations. During the late neolithic, gene pools across Anatolia and the Southern Caucasus mixed, resulting in an admixture cline^31^. The second event occurred during the Early Bronze Age, where Northern Levant populations, a region flanked by the Middle East and Europe, experienced gene flow in a process that likely involved a yet to-be-sampled neighboring population from Mesopotamia^31^. The most recent event for the modern Levant was largely determined by subsequent repopulations and mass movements associated with multiple cultural changes within the last two millennia. This appeared to have facilitated and maintained admixture between culturally different populations^32^. In general, the Jordanian population was not significantly different from their Levantine neighbours, and fit consistently into a Middle East-Anatolia-Balkan-Caucasus geographic and genetic continuum^33^.

On the other hand, correlation between *CYP2C9* genotypes and plasma levels of ibuprofen including level 1 evidence biomarkers with clinical pharmacogenetic guidelines^22^. These biomarkers were extensively studied for years and are already validated in clinical practice; a pharmacogenetic test prior to drug prescription is warranted. Mapped frequencies of these alleles showed that Central/South Asian, Near Eastern, and European populations were 7.9x more likely to show impaired *CYP2C9* metabolism than Sub-Saharan populations, and 4.9x more likely than East Asian ancestry populations (Table S7). Inference that a higher proportion of East Asian and African ancestry populations have normal ibuprofen metabolism, and therefore are less susceptible to complications for ibuprofen-based treatment. Figure 4 shows genotype frequencies of global populations of *CYP2C9*2* and *\*3* for more than 100,000 subjects within 412 reports^23^.

**Figure 4:**
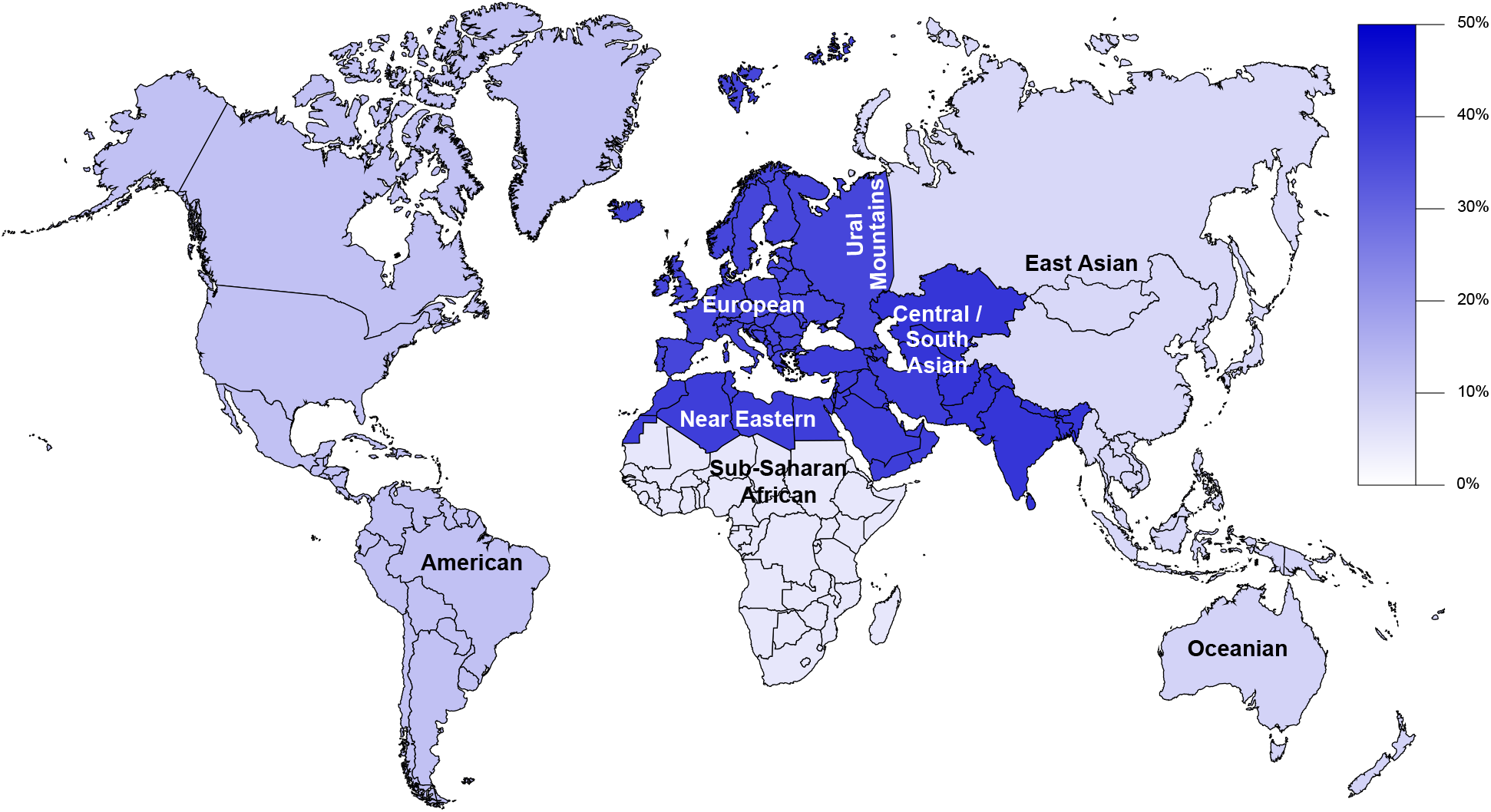
Global frequency map of variability in of *CYP2C9* level A alleles and impact on ibuprofen response. Phenotype prevalence shown for global populations of *CYP2C9*2* and *\*3*.

These findings are in agreement with several earlier European reports of potential harm with ibuprofen usage in patients with COVID-19 symptoms^4-9^. However, while the scientific debate whether to use or not ibuprofen in the course of COVID-19 continues^10-13^, three recent studies have recently been published with no significant association between ibuprofen prescription claims and severe COVID-19 in three countries, Saudi Arabia, Denmark and Israel, respectively^14-16^. These studies have several limitations. First, they were observational or retrospective studies; no causal conclusion can be reached, and recall-bias is a concern^14, 16^. Second, as those studies were conducted at an early stage of the pandemic outbreak, screening strategies in the beginning may have introduced selection bias relative to strategies at a later period^14-16^, such as, the policy of the Israeli Ministry of Health was to test only those individuals with symptoms that are suggestive of COVID-19. As a result, they have no information about ibuprofen in asymptomatic carriers, and whether antipyretics can influence their clinical course^16^. Third, the main analysis of these studies compared NSAID users with non-NSAID users, but confounding indicators may have influenced the results^14^, whereas ibuprofen users were defined based on prescription fills and information on whether or how many ibuprofen pills the patients actually took is not available^15^. Ibuprofen also can be purchased as an over-the-counter drug, they cannot exclude potential misclassification of ibuprofen exposure, as patients in the non-ibuprofen group may have used over-the-counter ibuprofen^15^. Lastly, their sample size was not sufficient to allow multivariable analyses^14, 16^.

However, the purpose of this study is not to confirm or deny the association between ibuprofen prescription claims and severe COVID-19. Alternatively, our simulation is based upon expected phenotype frequencies instead of directly assaying patient biospecimens, these phenotype frequencies are previously well-described, and are already validated in clinical practice^22, 23^. The adjusted expected phenotype frequencies based on the racial demographics of our cohort and expected population phenotype frequencies are dependent on population ancestry^34^.

Our analysis also revealed that other variants, such as, rs1057911 marker was found to be in LD with the variant of *CYP2C9*3* (*rs1057910*; Figure 1A, Table S2), which is consistent with the recently published PharmVar change adding *c*.*1425A>T* (*rs1057911*) to the *\*3* haplotype definition^35^. Further analysis indicated that the array probe used for genotyping may not able to bind specifically with the target SNPs, due to non-specific binding to another genomic region. Awareness of problematic regions is critical during test design and reporting to guide decisions regarding exclusion of regions and/or whether alternative assays must be used. This is particularly the case for *CYP2C9*7_5080C>A(L19I)* (*rs67807361*), where both statistical and genetic tests revealed a homologous sequence that may result in false positive or false negative variant calls.

A well-designed gene detection panel is important to fully characterize patient genotypes which relate to the drug effects. Although precision medicine should be considered in COVID-19 treatment, there are still many challenges to face. Firstly, a suitable strategy for treatment of COVID-19 through integrating pharmacogenomics data is still lacking. Secondly, the best treatment regime can be optimized for each individual patient, and both the efficacy and the safety can be guaranteed. Existing *CYP2C9* genotype results may provide the potential benefit of identifying populations who are at an increased risk of experiencing ADRs or therapeutic failure. This work demonstrates the capability and application of large-scale pharmacogenomics studies to elucidate genetic variation effects on treatment efficacy in COVID-19 patients. Ultimately, the implementation of pharmacogenetics in clinical settings is necessary as it leads to more efficient and cost-effective treatments.

## METHODS

### Sample collection

This study retrospectively included 101 unrelated Jordanian participants, of which 56 were male and 45 were female. After signed informed consent, a 3mL venous blood sample was collected in EDTA from each participant at the Princess Haya Biotechnology Centre between May 2010 and December 2011. Blood samples were stored at 4°C until DNA extraction. The Institutional Review Board (IRB) of the Jordan University of Science and Technology approved this study on 4/7/2013 under registration number 67/2/2013, and performed in accordance with the principles enshrined in the Declaration of Helsinki.

### DNA extraction and genotyping

Genomic DNA was extracted from each blood sample using the QIAamp DNA Micro Kit (Qiagen, Hilden, Germany) according to the manufacturer’s instructions. The quality of the purified DNA was determined using a NanoDrop ND-1000 spectrophotometer (Thermo Fisher Scientific, Waltham, MA USA).

Genotyping was accomplished using the Affymetrix DMET (Drug Metabolizing Enzymes and Transporters) Plus Premier microarray assay (Santa Clara, CA, USA) to test for drug metabolism associations. The DMET array contains 1,936 drug metabolism markers consisting of 1,931 single nucleotide polymorphisms (SNPs) and five copy number variations (CNVs) in 225 genes. These genetic variants were multiplex genotyped using molecular inversion probe (MIP) technology^29^. The profiles for the genotyping call rates and concordance comparisons, were generated by the DMET console software v1.3 (Thermo Fisher Scientific, Inc., Waltham, MA, USA), based on the Bayesian robust linear model with Mahalanobis (BRLMM) distance classifier algorithm^36, 37^. Genotypes were determined for each SNP site and reported as homozygous wild-type, heterozygous, homozygous variant, and ‘no call’ in the case of a lack of genotype call. SNPs with a call rate of less than 99% were excluded from subsequent analyses. Statistical and genetic analyses were performed for selection and validation using Microsoft Excel and SPSS v16. Linkage disequilibrium (LD) analysis was performed to identify non-random SNP associations between populations. LD was in concordance with all worldwide-distributed 1kG-p3 populations^38^. The genotype and allele frequencies were calculated and tested using the chi-square (χ^2^) test and the Hardy–Weinberg equilibrium formula (*p*□>□0.05) (Figure 5).

**Figure 5.**
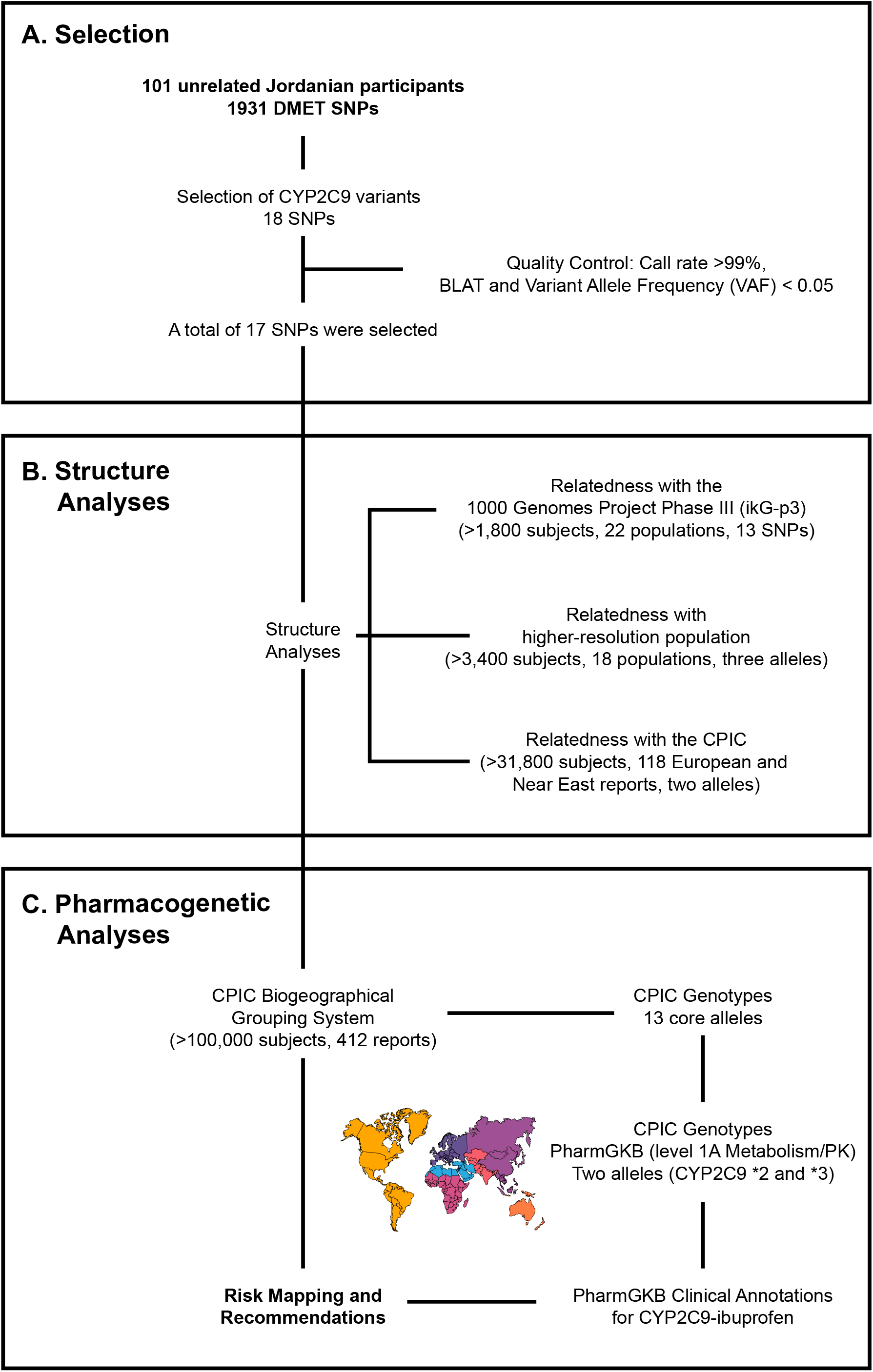
Flow chart depicting various steps involved in variant selection, analysis, and annotation. **a**. Selection of CYP2C9 variations by several statistical and genetic analyses. **b**. Structure analysis by multidimensional scaling (MDS) plots and fixation Index (Fst) was used to quantify population differentiation. **c**. Finally, the findings were adapted with nine geographically-defined groups (>100,000 subjects, 412 reports) in order to estimate genotype-predicted phenotype status across world populations.

### Selection of CYP2C9 variants

LD analysis was performed using the LDlink tool to generate D’ and r^2^ values^39^, and a matrix was generated for visualization. These allele frequencies were compared to 139 different CPIC reports from European and Near Eastern population groups (Table S8). The statistical comparison of allele frequencies on experimental data and reference populations were performed by Pearson’s χ^2^ test with Bonferroni correction and the negative logarithm of the adjusted significance values [-log10 (adj.p.val)] using the R statistical package v3.6.2 with ggplot2 and visualized using Rstudio v1.3.1056 (Boston, MA). The assembled DNA flanking sequence for each of the SNP loci was also subjected to BLAT^40^ to determine the specificity of the array’s probes matched to the target sequence set. A probe alignment was considered to be specific if 40 consecutive base pairs of the probe were fully aligned with the target sequence^29^. In order to detect the homologous sequences, which likely result in false-positive or false-negative variant calls, sequence similarity searches were performed using the Ensembl BLAST/BLAT search programs with default parameters.

### Population structure analyses

To identify cryptic relatedness from the genomic data, principal component analysis (PCA) was performed using the base R function “prcomp” within the R software package and multidimensional scaling (MDS) plots were generated from the PCA results using ggfortify and ggplot2^41^. Cryptic population structure was inferred using *CYP2C9* SNP data to identify the ancestral relatedness between the Jordanian Arab population and three defined datasets: 1,810 individuals from 22 populations from the 1000 Genomes Project Phase III (1kG-p3) dataset, excluding admixed populations (Table S9); 3,413 individuals from 18 global reports (Table S5); and 31,880 individuals from 118 reports from the European (EUR) and Near Eastern (NEA) populations from the CPIC updated report in March 2020 (Table S10).

Fixation Index (Fst) was used to quantify population differentiation from genetic structure using SNP allele frequencies. The R package “BEDASSLE” function was used to assess genetic similarity between ethnic populations by generating pairwise Fst values (0 indicates no divergence, 1 indicates complete separation) between the Jordanian Arab population and the other populations listed above^41^.

### Defined population datasets for structure analyses

Allele counts for defined populations were obtained from three sources, ethnic defined populations by the 1000 Genomes Project Phase III (1kG-p3) dataset^38^, consisting of 1,810 individuals from 22 populations within four defined ancestral groups (Table S9). The admixed populations were excluded to simplify the ethnic identification analyses.

1. African (AFR): Americans of African Ancestry in SW, USA (ASW), Esan in Nigeria (ESN), Gambian in Western Divisions in the Gambia (GWD), Mende in Sierra Leone (MSL), Luhya in Webuye, Kenya (LWK), Yoruba in Ibadan, Nigeria (YRI) and African Caribbeans in Barbados (ACB).
2. European (EUR): Utah Residents (CEPH) with Northern and Western European Ancestry (CEU), Finnish in Finland (FIN), British in England and Scotland (GBR), Iberian Population in Spain (IBS) and Toscani in Italia (TSI).
3. East Asian (EAS): Han Chinese in Beijing, China (CHB), Chinese Dai in Xishuangbanna, China (CDX), Southern Han Chinese (CHS), Japanese in Tokyo, Japan (JPT) and Kinh in Ho Chi Minh City, Vietnam (KHV).
4. South Asian (SAS): Bengali from Bangladesh (BEB), Indian Telugu from the UK (ITU), Punjabi from Lahore, Pakistan (PJL), Tamil from the UK (STU) and Gujarati Indian from Houston, Texas (GIH).

Two more higher-resolution groups of populations consisting of 3,413 individuals from 18 global reports were analyzed (Table S5). The findings were subsequently validated by increasing the coverage of this structure analysis to include more populations from the CPIC group across two geographically-defined groups: European (EUR) and Near Eastern (NEA). The genotypic data of individuals from 118 reports consisting of 31,880 individuals (Table S10), were downloaded from the CPIC updated report in March 2020 (Figure 5B).

### Biogeographic grouping system for Pharmacogenetic analyses

To standardize the reporting of ibuprofen susceptibility to develop complications by *CYP2C9* variability, nine groups consisting of seven geographically-defined groups (American, Central/South Asian, East Asian, European, Near Eastern, Oceanian, and Sub-Saharan African) and two admixed groups (African American/Afro-Caribbean and Latino) are presented in Figure 5C. These nine groups were defined by global autosomal genetic structure and based on data from large-scale sequencing initiatives^34^, are used to illustrate the broad diversity of global allele frequencies in this study^22, 23, 34, 42, 43, 44^. Furthermore, this biogeographic grouping system meets a key need in pharmacogenetic research by enabling consistent communication of the scale of variability in global allele frequencies and is now used by PharmGKB and CPIC^22, 23, 34^. The genotypic data of individuals from 412 global populations were downloaded from the CPIC updated report in March 2020.

1. American (AME): The American genetic ancestry group includes populations from both North and South America with ancestors predating European colonization, including American Indian, Alaska Native, First Nations, Inuit, and Métis in Canada, and Indigenous peoples of Central and South America.
2. Central/South Asian (SAS): The Central and South Asian genetic ancestry group includes populations from Pakistan, Sri Lanka, Bangladesh, India, and ranges from Afghanistan to the western border of China.
3. East Asian (EAS): The East Asian genetic ancestry group includes populations from Japan, Korea, and China, and stretches from mainland Southeast Asia through the islands of Southeast Asia. In addition, it includes portions of central Asia and Russia east of the Ural Mountains.
4. European (EUR): The European genetic ancestry group includes populations of primarily European descent, including European Americans. We define the European region as extending west from the Ural Mountains and south to the Turkish and Bulgarian border.
5. Near Eastern (NEA): The Near Eastern genetic ancestry group encompasses populations from northern Africa, the Middle East, and the Caucasus. It includes Turkey and African nations north of the Saharan Desert.
6. Oceanian (OCE): The Oceanian genetic ancestry group includes pre-colonial populations of the Pacific Islands, including Hawaii, Australia, and Papua New Guinea.
7. Sub-Saharan African (SSA): The Sub-Saharan African genetic ancestry group includes individuals from all regions in Sub-Saharan Africa, including Madagascar^8^.
8. African American/Afro-Caribbean (AAC): Individuals in the African American/Afro-Caribbean genetic ancestry group reflect the extensive admixture between African, European, and Indigenous ancestries and, as such, display a unique genetic profile compared to individuals from each of those lineages alone. Examples within this cluster include the Coriell Institute’s African Caribbean in Barbados (ACB) population and the African Americans from the Southwest US (ASW) population^9^ and individuals from Jamaica and the US Virgin Islands.
9. Latino (LAT): The Latino genetic ancestry group is not defined by an exclusive geographic region, but includes individuals of Mestizo descent, individuals from Latin America, and self-identified Latino individuals in the United States. Like the African American/Afro-Caribbean group, the admixture in this population creates a unique genetic pattern compared to any of the discrete geographic regions, with individuals reflecting mixed native and indigenous American, European, and African ancestry.

### Pharmacogenetic analyses

The frequencies of the 13 actionable pharmacogenomics biomarkers were assessed cumulatively for the Jordanian Arabs against nine biogeographical groups, consisting of 101,407 individuals from 412 global populations^23^. These nine groups were defined by global autosomal genetic structure and based on data from large-scale sequencing initiatives^34^ and are used to illustrate the broad diversity of global allele frequencies in this study. Furthermore,this biogeographic grouping system meets a key need in pharmacogenetics research by enabling consistent communication of the scale of variability in global allele frequencies and are now used by PharmGKB and CPIC^22^.

The total frequency of two SNPs with a Level 1A for the Jordanian Arab population within nine geographically-defined groups were mapped for global impact visualization of allele frequency on ibuprofen response (Table 3, Figure 5). Inferred frequency for CYP2C9*1 was excluded from our biogeographical analyses as no population studies have tested for all known variant alleles, and *\*1* was not genotyped directly in many studies^23^.

## Data Availability

The datasets generated during the current study are included in the supplementary files.

https://api.pharmgkb.org/v1/download/file/attachment/CYP2C9_frequency_table.xlsx

https://api.pharmgkb.org/v1/download/file/attachment/CYP2C9_Diplotype_Phenotype_Table.xlsx

## ACKNOWLEDGEMENTS

The technical staff from Princess Haya Biotechnology Center at the Jordan University of Science and Technology provided their assistance with this study.

## AUTHORS’ CONTRIBUTIONS

AA conceived the research study, performed all the analyses reported and wrote the manuscript.

## CONSENT FOR PUBLICATION

Consent for publication of the data obtained in this study was retrospectively approved, following de-identifying information of individuals.

## DATA AVAILABILITY

The datasets generated during the current study are included in the supplementary files. Publicly available repositories used in the analyses were previously published under PMID: 32189324 and downloaded from https://api.pharmgkb.org/v1/download/file/attachment/CYP2C9_frequency_table.xlsx, and https://api.pharmgkb.org/v1/download/file/attachment/CYP2C9_Diplotype_Phenotype_Table.xlsx.

## ETHICS DECLARATIONS

Signed informed consent was obtained from all participants recruited for this study under the ethical approval from University Review Committee for Research on Humans at Jordan University of Science and Technology (JUST) registered under [67/2/2013] on 4/7/2013, and performed in accordance with the principles enshrined in the Declaration of Helsinki.

## References

[1] Zhu, N. et al.. A Novel Coronavirus from Patients with Pneumonia in China, 2019. N Engl J Med. 382, 727–733 (2020).

[2] Lin, L. & Li, T. S. Interpretation of “Guidelines for the Diagnosis and Treatment of Novel Coronavirus (2019-nCoV) Infection by the National Health Commission (Trial Version 5)”. Zhonghua Yi Xue Za Zhi. 100, 805–807 (2020).

[3] Fang, L., Karakiulakis, G., & Roth, M. Are patients with hypertension and diabetes mellitus at increased risk for COVID-19 infection?. The Lancet. Respiratory Medicine. 8, e21. https://doi.org/10.1016/S2213-2600(20)30116-8 (2020).

[4] ANSM. COVID-19: the ANSM takes measures to promote the proper use of paracetamol updated on 17/03/2020. 2020. https://ansm.sante.fr/S-informer/Points-d-information-Points-d-information/COVID-19-l-ANSM-prend-des-mesures-pour-favoriser-le-bon-usage-du-paracetamol (2020).

[5] Little, P. Non-steroidal anti-inflammatory drugs and covid-19. BMJ. 368, m1185 (2020).

[6] Day, M. Covid-19: ibuprofen should not be used for managing symptoms, say doctors and scientists. BMJ. 368, m1086 (2020).

[7] Day, M. Covid-19: European drugs agency to review safety of ibuprofen. BMJ. 368, m1168 (2020)

[8] ANSM. COVID-19: Use of medicines in cities during the Covid-19 epidemic: update after five weeks of confinement - Information point updated on 05/04/2020. https://www.ansm.sante.fr/S-informer/Points-d-information-Points-d-information/Usage-des-medicaments-en-ville-durant-l-epidemie-de-Covid-19-point-de-situation-apres-cinq-semaines-de-confinement-Point-d-information (2020).

[9] Medicines and Healthcare products Regulatory Agency. Ibuprofen use and Coronavirus (COVID-19). https://www.gov.uk/government/news/ibuprofen-use-and-covid19coronavirus (2020).

[10] Moore, N., Carleton, B., Blin, P., Bosco-Levy, P., & Droz, C. Does Ibuprofen Worsen COVID-19? Drug Safety. 43(7), 611–614 (2020).

[11] Vosu, J. et al.. Is the risk of ibuprofen or other non-steroidal anti-inflammatory drugs increased in COVID-19? J Paediatr Child Health. 56, 1645–6 (2020).

[12] Kutti Sridharan, G. et al. COVID-19 and avoiding ibuprofen. How good is the evidence? Am J Ther. 27, e400–2 (2020).

[13] Sodhi, M. & Etminan, M. Safety of ibuprofen in patients with COVID-19: causal or confounded? Chest, 158. 55–6 (2020).

[14] Abu Esba, L. C. et al. Ibuprofen and NSAID use in COVID-19 infected patients is not associated with worse outcomes: a prospective cohort study. Infect Dis Ther. 2, 1–16 (2020).

[15] Kragholm, K. et al. Association Between Prescribed Ibuprofen and Severe COVID-19 Infection: A Nationwide Register-Based Cohort Study. Clin Transl Sci. 13. 1103– 1107 (2020).

[16] Rinott, E., Kozer, E., Shapira, Y., Bar-Haim, A. & Youngster, I. Ibuprofen use and clinical outcomes in COVID-19 patients. Clin Microbiol Infect. 26, 1259.e5–7.7 (2020)

[17] Gray, J. A. The shift to personalised and population medicine. Lancet. 382, 200–201 (2013).

[18] Peterson, J. F. et al. Building evidence and measuring clinical outcomes for genomic medicine. Lancet. 394, 604–610 (2019).

[19] Burchard, E. G. et al. The importance of race and ethnic background in biomedical research and clinical practice. N Engl J Med. 348, 1170–1175 (2003).

[20] Gemmati, D. & Tisato, V. Genetic Hypothesis and Pharmacogenetics Side of Renin-Angiotensin-System in COVID-19. Genes (Basel). 11, 1044 (2020).

[21] Ramamoorthy, A., Pacanowski, M. A., Bull, J. & Zhang, L. Racial/ethnic differences in drug disposition and response: review of recently approved drugs. Clin Pharmacol Ther. 97, 263–273 (2015).

[22] Whirl-Carrillo, M. et al. Pharmacogenomics knowledge for personalized medicine. Clin Pharmacol Ther. 92, 414–417 (2012).

[23] Theken, K. N. et al. Clinical Pharmacogenetics Implementation Consortium Guideline (CPIC) for CYP2C9 and Nonsteroidal Anti-Inflammatory Drugs. Clin Pharmacol Ther. 108, 191–200 (2020).

[24] Garcia-Martin, E., Martinez, C., Tabares, B., Frias, J. & Agundez, J. A. Interindividual variability in ibuprofen pharmacokinetics is related to interaction of cytochrome P450 2C8 and 2C9 amino acid polymorphisms. Clin Pharmacol Ther. 76, 119–127 (2004).

[25] Karazniewicz-Lada, M., Luczak, M. & Glowka, F. Pharmacokinetic studies of enantiomers of ibuprofen and its chiral metabolites in humans with different variants of genes coding CYP2C8 and CYP2C9 isoenzymes. Xenobiotica. 39, 476–485 (2009).

[26] Kirchheiner, J. et al. Enantiospecific effects of cytochrome P450 2C9 amino acid variants on ibuprofen pharmacokinetics and on the inhibition of cyclooxygenases 1 and 2. Clin Pharmacol Ther. 72, 62–75 (2002).

[27] López-Rodríguez, R. et al. Influence of CYP2C8 and CYP2C9 polymorphisms on pharmacokinetic and pharmacodynamic parameters of racemic and enantiomeric forms of ibuprofen in healthy volunteers. Pharmacol Res. 58, 77–84 (2008).

[28] Ochoa, D. et al. Effect of gender and CYP2C9 and CYP2C8 polymorphisms on the pharmacokinetics of ibuprofen enantiomers. Pharmacogenomics. 16: 939–948 (2015).

[29] Wang, Y., Cottman, M. & Schiffman, J. D. Molecular inversion probes: a novel microarray technology and its application in cancer research. Cancer Genet. 205, 341–355 (2012).

[30] Haber, M. et al. Genome-wide diversity in the levant reveals recent structuring by culture. PLoS Genet. 9, e1003316 (2013).

[31] Skourtanioti, E. et al. Genomic History of Neolithic to Bronze Age Anatolia, Northern Levant, and Southern Caucasus. Cell. 181, 1158–1175 (2020).

[32] Scozzari, R. et al. Human Y-chromosome variation in the western Mediterranean area: implications for the peopling of the region. Hum Immunol. 62, 871–884 (2001).

[33] Flores, C. et al. Isolates in a corridor of migrations: a high-resolution analysis of Y-chromosome variation in Jordan. J Hum Genet. 50, 435–441 (2005).

[34] Huddart, R. et al. Standardized Biogeographic Grouping System for Annotating Populations in Pharmacogenetic Research. Clin Pharmacol Ther. 105, 1256–1262 (2019).

[35] Gaedigk, A. et al. The Pharmacogene Variation (PharmVar) Consortium: Incorporation of the Human Cytochrome P450 (CYP) Allele Nomenclature Database. Clin Pharmacol Ther. 103, 399–401 (2018).

[36] Rabbee, N. & Speed, T. P. A genotype calling algorithm for affymetrix SNP arrays. Bioinformatics. 22, 7–12 (2006).

[37] Sissung, T. M., English, B. C., Venzon, D., Figg, W. D. & Deeken, J. F. Clinical pharmacology and pharmacogenetics in a genomics era: the DMET platform. Pharmacogenomics. 11, 89–103 (2010).

[38] 1000 Genomes Project Consortium., et al. A global reference for human genetic variation. Nature. 526, 68–74 (2015).

[39] Machiela, M. J. & Chanock, S. J. LD link: a web-based application for exploring population-specific haplotype structure and linking correlated alleles of possible functional variants. Bioinformatics. 31, 3555–3557 (2015).

[40] Kent, W. J. BLAT--the BLAST-like alignment tool. Genome Res. 12, 656–664 (2002).

[41] R Core Team. R: A language and environment for statistical computing. R Foundation for Statistical Computing. https://www.r-project.org (2013).

[42] Elhaik, E. et al. Geographic population structure analysis of worldwide human populations infers their biogeographical origins. Nat Commun. 5, 3513 (2014).

[43] Jakobsson, M. et al. Genotype, haplotype and copy-number variation in worldwide human populations. Nature. 451, 998–1003 (2008).

[44] Li, J. Z. et al. Worldwide human relationships inferred from genome-wide patterns of variation. Science. 319, 1100–1104 (2008).

